# Breast Cancer Segmentation and Diagnosis in Mammography: Toward a Pan-European Virtual Biopsy Framework

**DOI:** 10.1101/2025.05.09.25327297

**Authors:** Manuel Marfil-Trujillo, Aikaterini Vraka, Leonor Cerdá-Alberich, Gloria Ribas-Despuig, Silvia Flor-Arnal, Paula Jiménez-Gómez, Luis Martí-Bonmatí

## Abstract

**Background:** Most mammographic AI stops at a binary cancer decision and ignores the histopathologic information that guides therapy.

**Purpose:** To build an end-to-end virtual-biopsy pipeline that simultaneously segments tumours and predicts pT, pN, cM and molecular subtype across scanners and countries.

**Materials and Methods:** A residual U-Net was trained on 8 469 public mammograms and externally tested on 200 cases from six ChAImeleon sites. From each mask we derived 1 559 radiomics, 5 120 RadImageNet embeddings and 12 clinical covariates; a 5 × 3 nested cross-validation tuned an RBF-SVM/XGBoost/MLP ensemble with a random-forest meta-learner.

**Results:** External Dice was 0.458; adding benign masks cut false-positive pixels by 19% but left Dice unchanged (0.455).

## Introduction

Breast cancer is the most frequently diagnosed malignancy among women, with 2.3 million new cases in 2020 and a projected 47.8% increase by 2040 among women aged 15–49 years.[1] Although organised mammography screening has reduced mortality, deaths decline more slowly than incidence, revealing shortcomings in current workflows: over-diagnosis of low-risk lesions, substantial inter-reader variability (Cohen *κ <* 0.5 for subtle findings) and an escalating workload amid a shortage of breast radiologists.[2]

Precise tumour outlining is therefore the linchpin of any downstream “virtual-biopsy” pipeline. Yet most segmentation studies remain monocentric or rely on small, legacy repositories. For instance, the Indian DMID series (510 mammograms) was captured on a single Hologic detector, offering no vendor or ethnic diversity.[3] A systematic review of 1999–2021 methods found that *>* 70% of papers continue to train exclusively on MIAS (*n*=322) or INbreast (*n*=410) and seldom perform external testing.[4] Even the million-image NYU “GLAM” saliency dataset originates from one health-system,[5] while newer CESM-U-Net[6] and multi-scale graph approaches[7] report high Dice on *<* 100 cases, again from single centres. Such constraints inflate apparent accuracy yet mask *domain-shift* fragility: changes in detector model, reconstruction kernel, breast density or population mix often degrade performance. Robust segmentation therefore demands **multicentric, multivendor, richly annotated cohorts**, withholding entire sites—not random images—for external validation.

Imaging-based “virtual biopsy” has likewise advanced from coarse lesion typing to receptor-status prediction, but remains lesion-centric. Barros *et al.*[8] combined multimodal images from two vendor-matched sites to separate invasive, in-situ and benign disease. Zhang *et al.*[9] trained a supervised network on single-centre mammograms to predict combined ER/PR expression. A later dual-centre study inferred individual HER2, ER and PR status.[10] Nevertheless, no published work has yet demonstrated a virtual-biopsy model that concurrently predicts cellular subtype *and* nodal (N) as well as distant-metastatic (M) categories using truly multicentric woman cohorts—leaving this crucial diagnostic space unexplored.

**ChAImeleon**—a GDPR-compliant, pan-European oncology imaging biobank—contains *>* 2,500 breast-cancer cases collected in Spain, Italy, Portugal, Turkey, Lithuania and Croatia. Each record couples two-view digital mammograms (or synthetic 2-D reconstructions from DBT) with an OMOP-harmonised clinicopathologic profile: histotype, ER/PR/HER2/Ki-67, Nottingham grade, TNM stage, treatment timelines and outcomes. Images span four major vendors (Hologic, GE, Siemens, Fujifilm). This uniquely multicentric, multivendor and richly annotated repository provides the solid foundation for the virtual-biopsy framework proposed herein.

We hypothesised that coupling a robust public-dataset segmentation network with multimodal feature integration would deliver a non-invasive *virtual biopsy* capable of (i) precisely localising malignancies and (ii) predicting TNM stage and molecular subtype. Our aims were to (1) describe the end-to-end curation and preprocessing pipeline that minimises scanner/country domain shift; (2) externally validate segmentation on a stratified ChAImeleon subset; and (3) benchmark the stacked-ensemble classifier against prior literature, evaluating its potential to reduce unnecessary biopsies and guide personalised therapy.

## Materials and Methods

### Study Design

Retrospective, multi-institutional study compliant with the Declaration of Helsinki. Review boards of all contributing centres approved data reuse; written informed consent was waived for anonymised retrospective data.

### Training Data: Public Repositories

Seven public repositories—CBIS-DDSM[11], MIAS[12], INbreast[13], TOMPEI-CMMD[14], CSAW-CC[15], VinDr-Mammo[16] and EMBED[17]—were screened for two-view digital mammograms that contained at least one region of interest (ROI) classified as malignant. Dataset characteristics and licensing terms are summarised in Table 1, adapted from Logan *et al*.[18] A ROI was deemed *malignant* when the lesion had histopathologic confirmation (biopsy or surgical specimen); if pathology was unavailable, radiologic consensus of three independent readers rating BI-RADS *≥* 4 was accepted. All other annotated findings (BI-RADS *≤* 3) were labelled *benign* and used only as background masks. Curation was performed with Python 3.12:

**Table 1.**
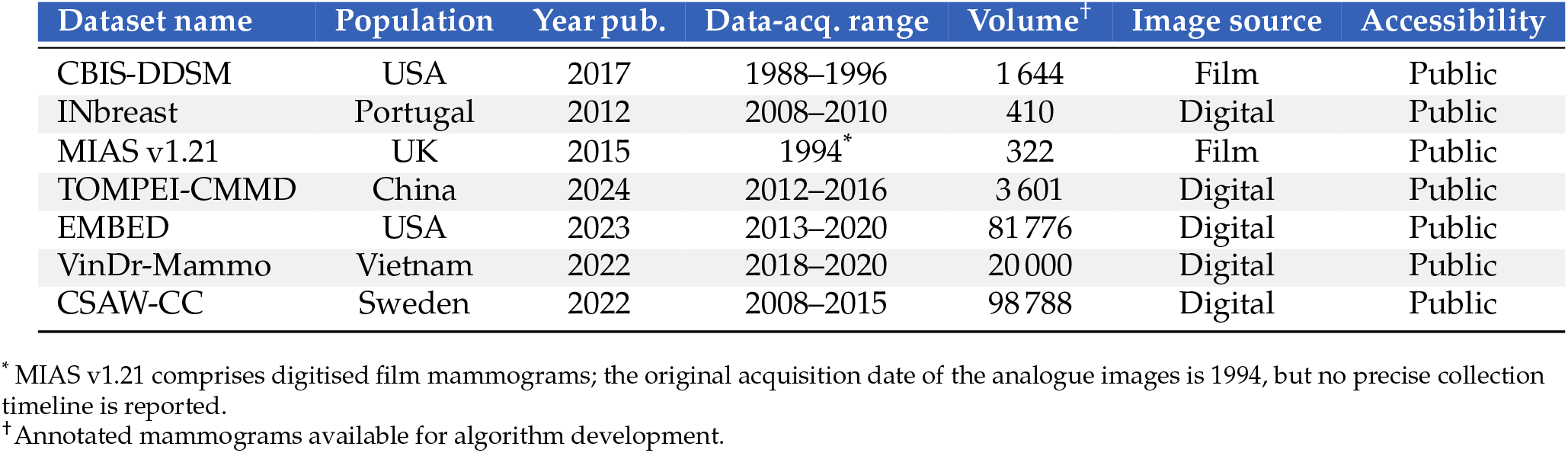
Public mammography repositories used for training (summary characteristics).

**Table 2.** Technical and annotation properties of the public training datasets.

| Dataset | Image format | ROI format | Diagnostic confirmation |
| --- | --- | --- | --- |
| CBIS-DDSM | DICOM | Binary mask & BBox | Histological |
| INbreast | DICOM | Polygon coordinates | Histological |
| MIAS v1.21 | PGM | Centre/radius coordinates | Histological |
| TOMPEI-CMMD | DICOM | Polygon coordinates | Histological |
| EMBED | DICOM | Bounding-box coordinates | Histological |
| VinDr-Mammo | DICOM | Bounding-box coordinates | Radiological<sup>1</sup> |
| CSAW-CC | DICOM | Binary mask | Radiological<sup>1</sup> |

<sup>1</sup> Two experienced breast radiologists reached consensus; if discrepant, a third senior reader adjudicated the final label.

**Table 3.** Breast-cancer cases available in the CHAIMELEON biobank, aggregated by country.

| Country | Breast-cancer cases |
| --- | --- |
| Spain | 1 297 |
| Italy | 526 |
| Portugal | 192 |
| Turkey | 264 |
| Lithuania | 235 |
| Croatia | 128 |

1. **Format conversion** to NIfTI while preserving spatial metadata: DICOM *→* NIfTI via `dcm2niix`v1.0.20240202 PGM *→* NIfTI via `imageio`v2.37 PNG *→* NIfTI via `SimpleITK`v2.4.1
2. **Mask generation** for datasets that supplied coordinates but no binary labels:
  - *Bounding-box ROIs*: rectangles were rasterised on zero arrays, copied DM spatial header and smoothed to an ellipse to avoid inclusion of healthy tissue.
  - *Polygon ROIs*: vertex lists in XML/JSON were filled with `scikit-image`v0.25.2.
  - *Circular ROIs*: centre and radius specified in XLSX were converted to distance maps using Numpy v1.26.4; pixels within the radius were set to 1.
3. **ROI fusion**: all malignant ROIs belonging to the same mammogram were merged into a single label map.
4. **Background definition**: benign ROIs were relabelled as background; mammograms containing exclusively benign findings were retained with an empty mask, while normal studies without annotations were excluded.

The final corpus comprised 8 730 DM–mask pairs: 4 433 masks with malignant ROIs and 4 297 empty masks containing only benign background.

### External Validation Cohort

From all cases stored in the ChAImeleon breast repository, an external-validation set of 200 cases was drawn by stratified random sampling. First, the stage-at-diagnosis distribution for each of the six contributing countries was retrieved from the most recent population-based registries and national audits.[19, 20] These country-specific proportions were combined as a weighted average, where each weight equalled that nation’s share of ChAImeleon cases; the resulting sampling quotas produced a cohort composition of *Stage I 44.0 %, Stage II 38.9 %, Stage III 15.3 % and Stage IV 1.8 %*.—closely reflecting real-world epidemiology. Four fellowship-trained breast radiologists (8–24 years of experience) independently delineated all malignant foci in 3D Slicer v5.6.2; disagreements were resolved in consensus sessions, producing the reference masks used for external validation.

### Image Pre-processing

Each digital mammogram was denoised with an adaptive median filter (initial window 3 × 3, maximum 7×7; Numba–optimised) and contrast-enhanced with CLAHE (clip_limit 0.01, 256 bins) [21, 22]. The enhanced 2-D volumes were then resampled to 1 mm isotropic spacing (cubic interpolation for images, linear for labels) and Z-score normalised channel-wise. Patch dimensions and normalisation statistics differed across different training set combinations and are reported in Table 4.

**Table 4.**
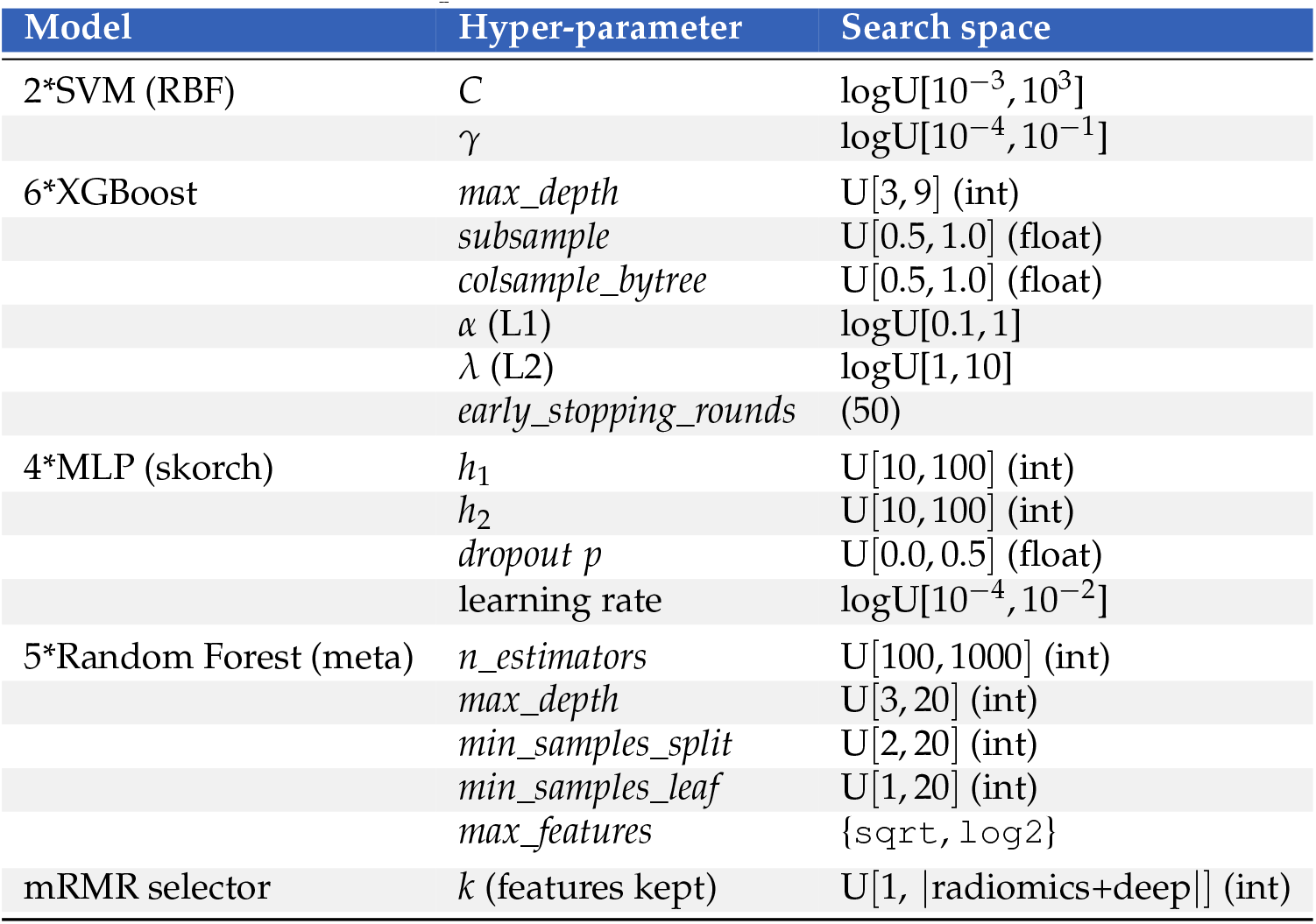
Search space explored in the *inner* loops of the nested CV. All trials used the Optuna TPE sampler (250 trials per base learner, 250 for the meta-learner). logU[*a, b*] denotes a log-uniform prior between *a* and *b*; U[*a, b*] a linear uniform prior; fixed values are indicated in parentheses.

### Segmentation Model Development

A nine-stage residual encoder U-Net with 3 × 3 convolutions per stage and 32–512 feature channels (InstanceNorm 2-D, Leaky-ReLU) was adopted. The largest anisotropic patch size that fit in GPU memory *(batch size = 13)* was selected, ensuring efficient context capture without manual tuning [23].

Training followed three curricula: (i) 8 469 DM–mask pairs from public repositories; (ii) 4 433 malignant public pairs; (iii) 4 681 malignant pairs (4 433 public + 248 from ChAimeleon holdout test set). Curricula (i) and (ii) were used to assess cross-validation performance and to evaluate data-mismatch between public datasets versus ChAImeleon’s repository subsample, and to compare whether the inclusion of a large number of DM cases with benign anomalies along with empty masks improves performance by reducing false positives. Curriculum (iii) yielded the inference model for the ChAImeleon’s repository population. Training was performed with Dice loss and batch-wise Dice supervision, using a polynomial learning rate schedule decay of 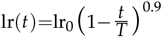where lr_0_ = 0.01, *t* is the current iteration and *T* is the total number of iterations. Five-fold cross-validation was executed in parallel on five NVIDIA A100-SXM4 80 GB GPUs. Folds were stopped early when validation loss plateaued for 50 epochs. During inference the soft-max outputs of the five folds were averaged and the ensemble mask binarised at 0.5.

**Figure 1.**
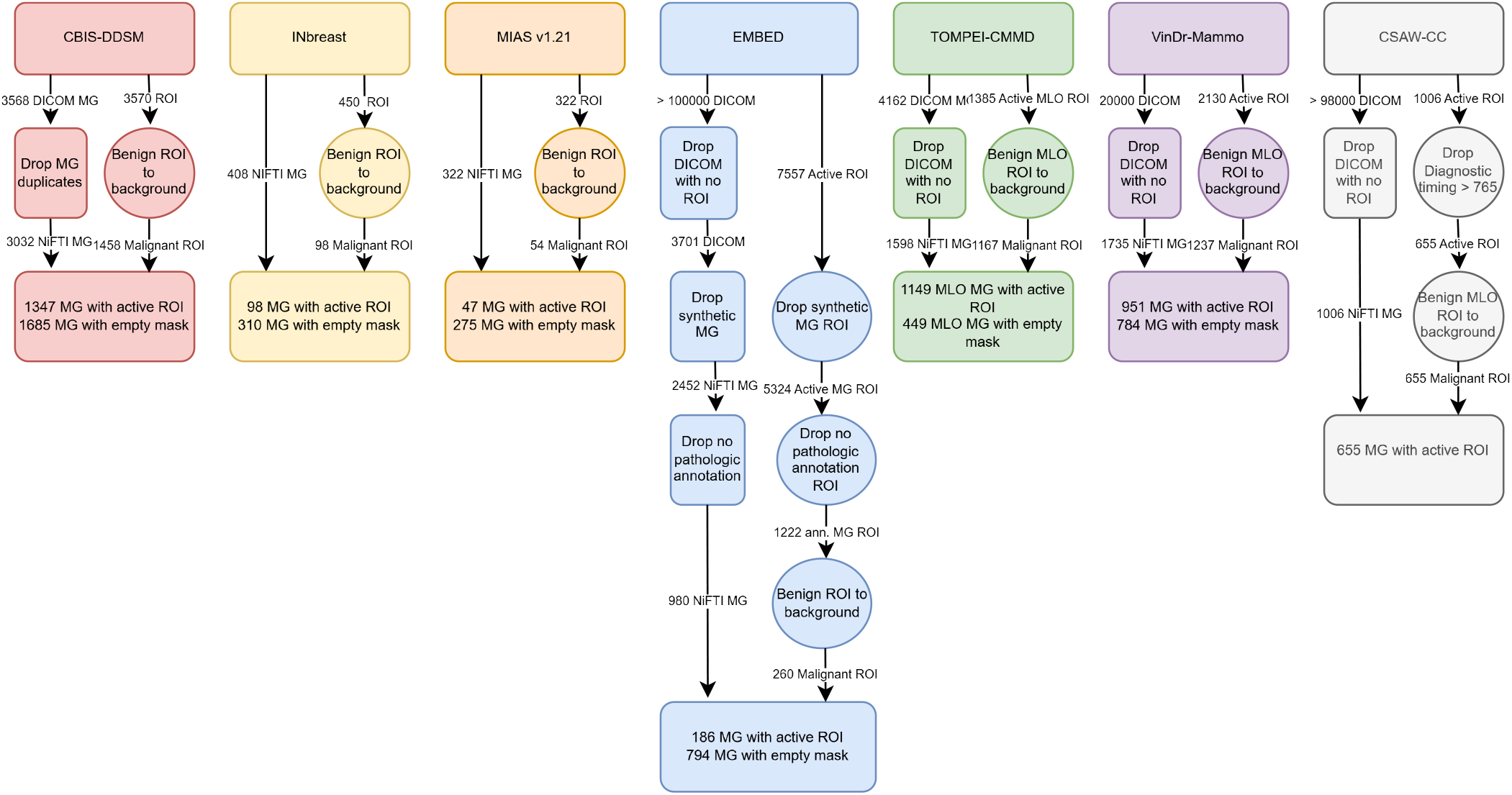
Overview of the data-curation, standardisation and fusion workflow.

### Inference Pipeline

Patients were eligible for inference only if at least one of the target clinical fields was present—tumour subtype, pathologic T (pT), pathologic N (pN) or clinical M (cM)—so that every subject contributed to at least one downstream model. For each eligible patient, all mammographic series were then screened and retained only when DICOM metadata met four image-quality criteria: Modality (0008, 0060)=`MG`, Presentation Intent Type (0008, 0068)=`FOR PRESENTATION`, Photometric Interpretation (0028, 0004)=`MONOCHROME2`and Image Laterality (0020, 0062)=`L/R`. Missing or ambiguous laterality tags were resolved by loading the pixel matrix, splitting it at mid-width and counting non-zero pixels in each half; the breast with the greater number of foreground pixels was assumed to contain tissue and therefore defined the laterality (right *>* left *→ R*, else *L*). View was identified from View Position (0018, 5101)=`CC/MLO/ML`. Clinical–pathologic data indicated the breast harbouring the malignant lesion, and from that side craniocaudal (CC) and mediolateral oblique (MLO) view were exported.

The final inference cohort comprised 3 781 pathology-confirmed malignant mammograms (TNM distribution 51.1 % Stage I, 31.4 % Stage II, 16.0 % Stage III, 1.5 % Stage IV). Prior to inference, the images underwent the identical preprocessing pipeline applied during training. Segmentation was subsequently performed with the previously trained U-Net ensemble.

### Radiomic Feature Extraction

Radiomics were extracted following the Image Biomarker Standardisation Initiative and ESR guidelines [24, 25]. The same DM pre-processing for inference (AMF denoising and CLAHE) were reused [26, 27]. Since data were multicentric, pixel intensities were *Z*-score normalised and values outside *±* 2.33*σ* (1st–99th percentiles) were clipped (`normalize=True`, `removeOutliers=2.33`, `normalizeScale=1`, `voxelArrayShift=0`). Each tumour mask was B-spline resampled to an isotropic grid of 0.062 mm × 0.062 mm,i.e. the smallest native spacing observed (`padDistance=5`, `interpolator=sitkBSpline`, `resampledPixelSpacing=[0.062, 0.062]`).

A cohort-wide scan of normalised ROI intensities established a 16–128 bin-count interval; its 25^th^, 50^th^ and 75^th^ percentiles were converted to three fixed bin-widths—0.011, 0.0035 and 0.0021—which yielded mean ± SD bin counts of 33.2± 2.1, 70.5± 5.2 and 101.8± 8.7 bins respectively. For every bin width, 2-D radiomics were extracted from nine `imageType`s (`Original`, `LoG`, `Wavelet`, `Square`, `SquareRoot`, `Logarithm`, `Exponential`, `Gradient`, `LocalBinaryPattern2D`) and all seven `featureClass`es (`FirstOrder`, `Shape2D`, `GLCM`, `GLRLM`, `GLSZM`, `GLDM`, `NGTDM`), producing 1 559 radiomic features. To select the most reproducible discretisation, each matrix was evaluated by computing absolute-agreement ICC_2_ and the mean per-patient coefficient of variation across replicate measurements; the bin-width configuration with the highest median ICC and lowest median CV was retained for downstream modelling.

**Figure 2.**
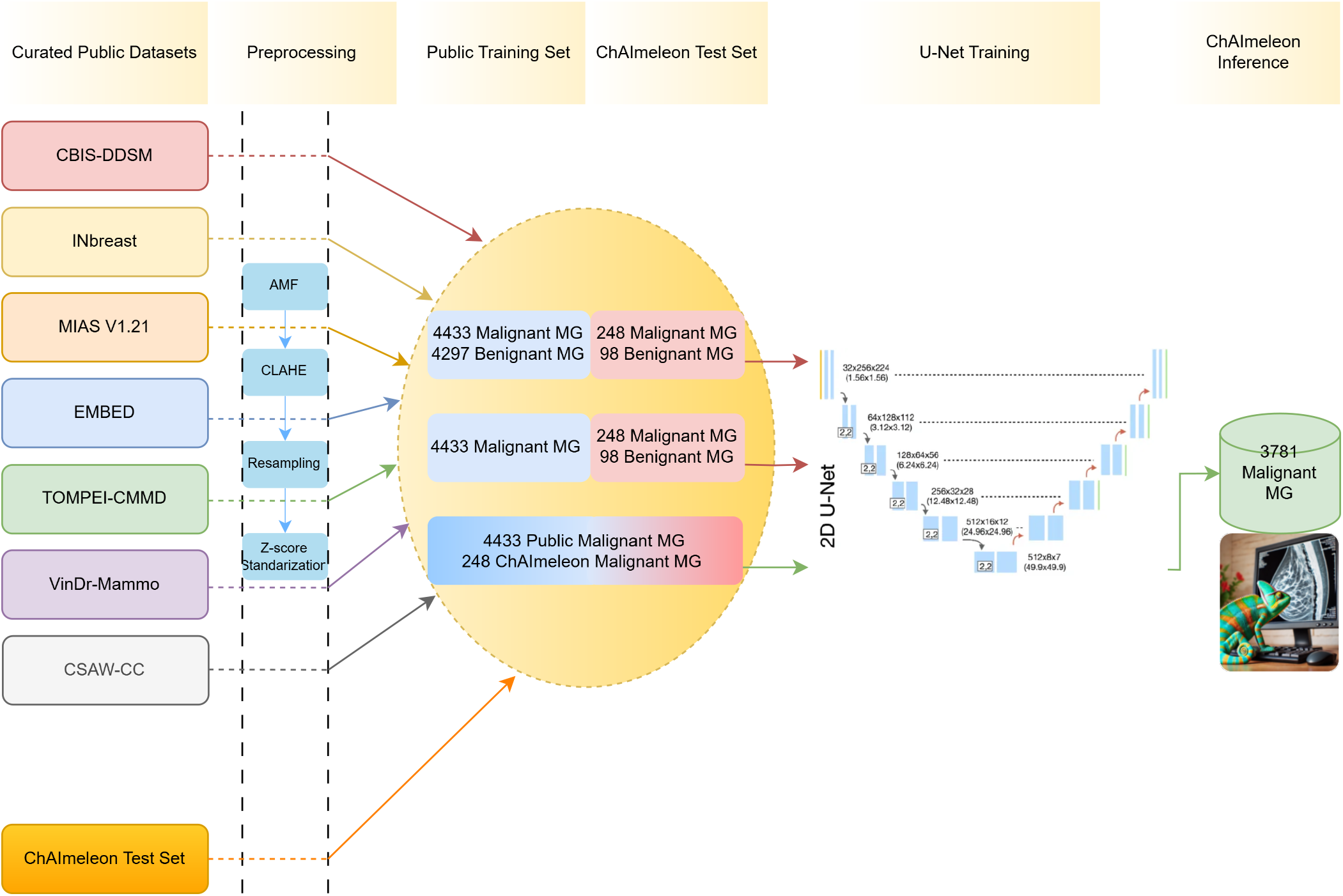
Pipeline for training, validating and ensemble-averaging the residual U-Net segmentation model.

### Deep Feature Extraction

After same pre-processing than for radiomics extraction (isotropic B-spline resampling to the smallest inplane pixel [0.062×0.062] and the Z-score normalization with 1st–99th percentiles clipped), the tightest bounding box enclosing each connected-component of the binary mask was created and its coordinates were used to crop the ROI of the DM. The patchs were encoded as 8-bit, one-channel and resized to 224 × 224 px for `ResNet-50`and `DenseNet-121`, or 299 × 299 px for `Inception-V3`[28, 29]. Feature vectors were read from the global-average-pooling layer of three trunks pretrained on RadImageNet, a public dataset of *~* 1.35 million radiologic images from multiple modalities. The resulting dimensionalities were 2 048 (`ResNet-50`), 1 024 (`DenseNet-121`) and 2 048 (`Inception-V3`).

### Clinical Variables

Twelve pre-biopsy variables recorded in the ChAImeleon eCRF were considered: *age at diagnosis, menopausal status, family history breast cancer* and *lesion position* within the breast quadrant map (*LIQ, LOQ, UIQ, UOQ, axillary, nipple, central*). *Scanner manufacturer* was included as a technical covariate.

### Pre-processing and model training

#### Tabular pre-processing

For each prediction target—histologic subtype (*apocrine, ductal-in situ, infiltrating ductal, lobular, mucinous, papillary, tubular*), tumour stage (*pT*_is_, *pT*0–*pT*4), nodal stage (*pN*0–*pN*3) and distant metastasis (*cM*0 vs. *cM*1)—the ChAImeleon cohort was stratified into an 80 % training and 20 % hold-out test split. Every pre-processing operator was fitted on each feature block of the training split and then transformed on the corresponding block of the test split to avoid data leakage.

The tabular features were organised in five blocks—radiomics, ResNet-50, DenseNet-121, Inception-V3 embeddings and clinical covariates—and *every* pre-processing operator listed below was fitted independently within each numeric block (clinical variables were exempt from ComBat and variance filtering). The transformed blocks were concatenated only after the last step to build the modelling matrix.

##### Missing-value imputation

numerical variables with a Bayesian-ridge `IterativeImputer`; categorical variables with `MissForest`.

##### Batch-effect correction

manufacturer effect removed per feature block with empirical-Bayes ComBat.

##### Variance filtering

within each numeric block the top 10% most-variable descriptors were retained.

##### Robust scaling

features were median-centred and scaled by their inter-quartile range.

#### Nested cross-validation and feature selection

Model development used a 5×3 nested cross-validation: the five outer folds provided an unbiased generalisation estimate, whereas each inner three-fold loop performed Bayesian hyper-parameter optimisation (250 trials, macro-F_1_ as the objective). All hyper-parameters explored are detailed in Table 4. Within every inner loop an additional step was executed:

##### mRMR selection

minimum redundancy maximum relevance (`mRMR`) was applied only to the concatenated radiomic and deep-embedding feature blocks; the top *k* features—where *k* was tuned during optimisation—were then appended to the complete set of clinical variables to form the final modelling matrix.

#### Ensemble architecture

For each outer fold the inner CV returned the best configuration for the three base learners and for the random-forest meta-learner. Base models were refit on the corresponding outer-training split; at inference their class-probability vectors were averaged across the five outer folds to create a meta-dataset, which was ingested by the five fold-specific meta-models. The final prediction was the mean of those five meta-model outputs.

Per-class sensitivity, specificity, precision, F_1_ and balanced accuracy were reported. Model interpretability was assessed by averaging SHAP values across the five outer meta-models and visualising them as class-specific summary plots.

## Results

### Tumor–ROI detection and segmentation

Despite outperforming its counterpart in internal fivefold cross-validation (Dice_cv_=0.671 vs 0.554), the lesion-only curriculum ultimately achieved almost the same external accuracy as the model trained with additional empty-mask images (Dice_test_=0.458 vs 0.455). Introducing healthy-tissue examples cut false-positive pixels by 19% and boosted precision, but this was offset by a 3% rise in false negatives and a slight fall in sensitivity. The net Dice thus remained unchanged because the precision gain compensated for the recall loss; the drastic drop from cross-validation to external Dice almost certainly reflects *data mismatch*, as ChAImeleon comprises women from six European countries and four vendor platforms with heterogeneous acquisition protocols that differ markedly from the legacy public datasets used for training. Consequently, superior internal metrics do not guarantee real-world performance, and adding empty masks merely produces a more conservative network—preferable when reducing radiologist workload outweighs missing a minority of tumour voxels.

**Figure 3.**
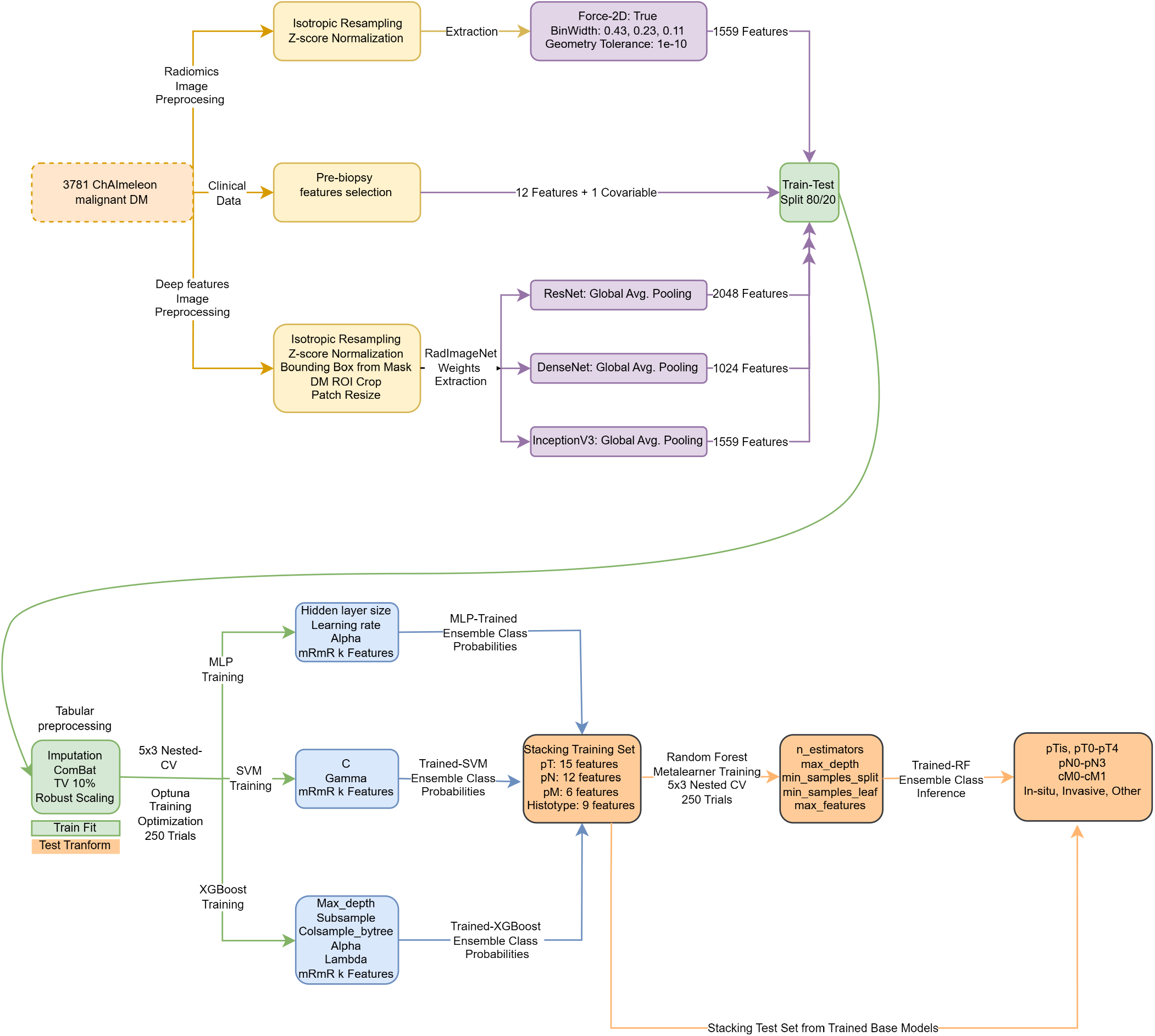
Complete workflow for variable harmonisation, feature selection and meta-learning based clinical prediction.

## Discussion

Our results confirm that cross-validation performance is a poor surrogate for multicenter, true generalizability in mammographic segmentation. Although Curriculum ii achieved a markedly higher internal Dice (0.671) than Curriculum i (0.554), both networks performed identically on the unseen ChAImeleon cohort (Dice = 0.46). This gap highlights the well-known *dataset-shift* between legacy public repositories, mainly single-vendor, film-digitized or North American DM, and the multicentric, multi-vendor European images in ChAImeleon. Adding empty benign masks (Curriculum 1) did not raise external Dice but almost halved false-positive pixels, suggesting that explicit exposure to normal anatomy teaches the model to suppress spurious tumour predictions. The modest increase in false-negatives (+3%) reflects the classical precision-recall trade-off and could be mitigated post-hoc by lowering the binarisation threshold or by cascade filtering with a high-sensitivity detector.

**Figure 4.**
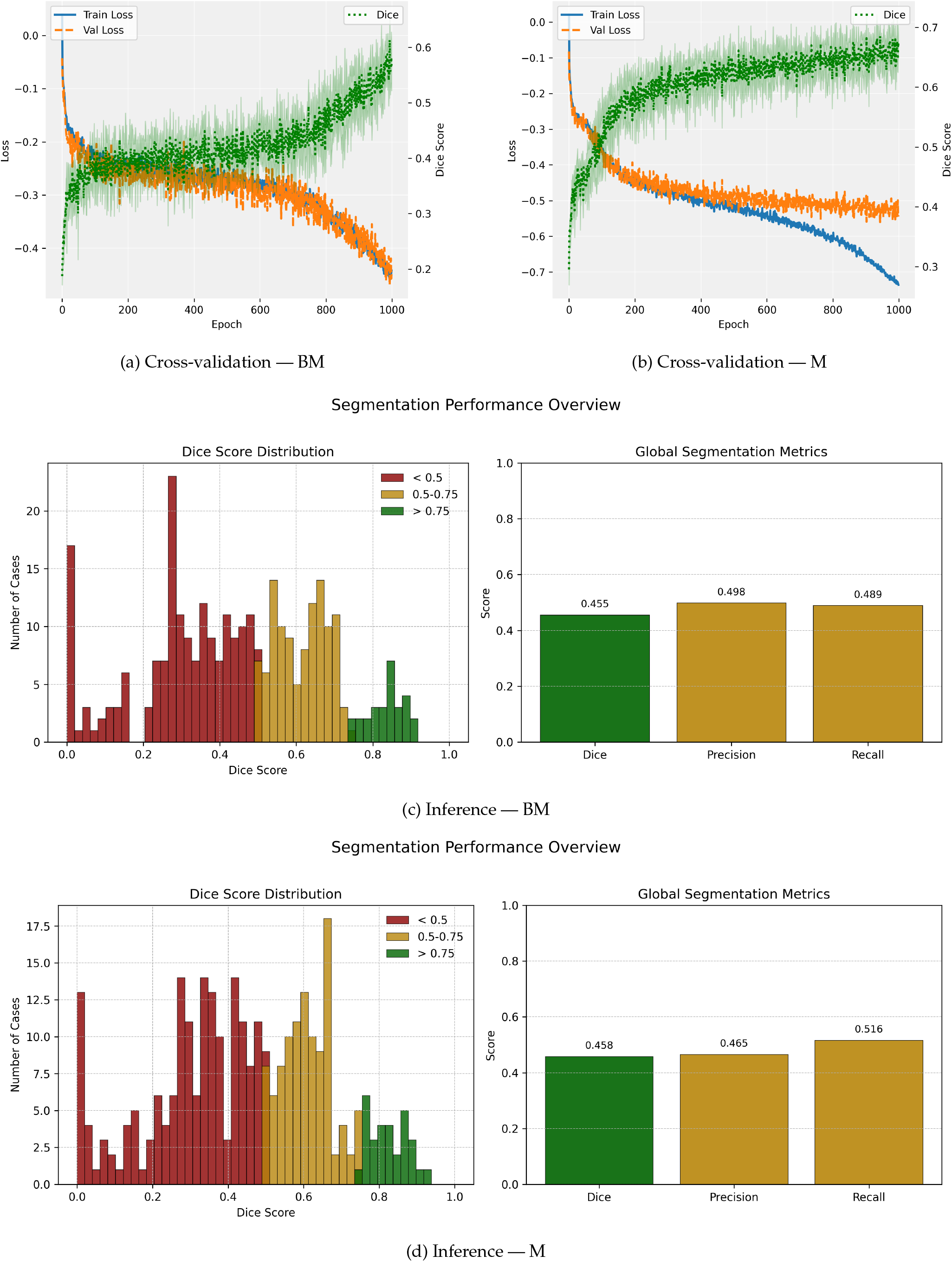
Performance overview of the two training curricula. **A**, five-fold cross-validation learning curve for Curriculum 1 (empty-mask + malignant ROIs). **B**, five-fold cross-validation learning curve for Curriculum 2 (malignant ROIs only). **C**, inference metrics of the Curriculum 1–trained model on the ChAImeleon test set. **D**, inference metrics of the Curriculum 2–trained model on the same test set.

## Data Availability

All data produced in the present study are available upon reasonable request to the authors

https://www.bing.com/search?qs=HS&pq=CBIS&sk=CSYN1HS1&sc=14-4&pglt=43&q=cbis-ddsm&cvid=636011fd469948589b88c27a5429facf&gs_lcrp=EgRlZGdlKgcIABAAGPkHMgcIABAAGPkHMgYIARAAGEAyBggCEAAYQDIGCAMQRRg5MgYIBBBFGDsyBggFEC4YQDIGCAYQABhAMgYIBxBFGDwyBggIEEUYPTIICAkQ6QcY_FXSAQgxNzE5ajBqMagCALACAA&FORM=ANNAB1&PC=U531

https://www.kaggle.com/datasets/martholi/inbreast

https://www.kaggle.com/datasets/kmader/mias-mammography

https://researchdata.se/en/catalogue/dataset/2021-204-1

https://www.bing.com/search?q=embed%20breast%20dataset&qs=n&form=QBRE&sp=-1&ghc=1&lq=0&pq=embed%20breast%20datase&sc=1-19&sk=&cvid=E51F41007FA04854A5059769F9C88280

https://www.bing.com/search?q=tompei+cmmd&cvid=3f1d2baa0a1b41f99ef0448b375e12ad&gs_lcrp=EgRlZGdlKgYIABBFGDkyBggAEEUYOTIGCAEQRRg8MgYIAhBFGD0yCAgDEOkHGPxV0gEIMTg4NWowajSoAgCwAgA&FORM=ANAB01&PC=U531

https://vindr.ai/datasets/mammo

## References

[1] Cai Y, Dai F, Ye Y, Qian J. The global burden of breast cancer among women of reproductive age: a comprehensive analysis. Sci Rep. 2025;15:9347.

[2] Ahsen ME, Ayvaci MUS, Mookerjee R, Stolovitzky G. Economics of AI and human task sharing for decision-making in screening mammography. Nat Commun. 2025;16:2289.

[3] Sarkar K, Banerjee P, Chaki N, et al. DMID: an Indian digital-mammography dataset for breast-lesion segmentation and classification. J Digit Imaging. 2023;36:1234–1246.

[4] Agarwal S, Tripathi S, Choudhary T, et al. Deep-learning approaches for breast-lesion segmentation: a comprehensive review of 1999–2021 literature. Biomed Res Int. 2021;2021:9962109.

[5] Wu N, Huang Y, Park J, et al. Deep neural networks improve radiologists’ performance in breast-cancer screening. IEEE Trans Med Imaging. 2019;38:1184–1194.

[6] Jiang Y, Wang L, Zhang Q, et al. CESM-U-Net: mass segmentation in contrast-enhanced spectral mammography using an improved U-Net. Comput Methods Programs Biomed. 2022;221:106882.

[7] Lin H, Zhou X, Wu J, et al. M3D-Graph: multi-scale 3-D graph convolution for automatic breast-tumour segmentation in digital mammograms. PLoS One. 2024;19:e0309421.

[8] Barros V, Tlusty T, Barkan E, et al. Virtual biopsy by using AI-based multimodal modelling of binational mammography data. Radiology. 2023;306:e220027.

[9] Zhang L, Li Y, Gao Y, et al. Weakly supervised deep learning of screening mammograms predicts combined oestrogen-progesterone receptor status. Sci Rep. 2023;13:17894.

[10] Zeng S, Chen H, Jing R, et al. Assessment of HER2, ER and PR expressions based on mammography using deep learning. Sci Rep. 2025;15:4826.

[11] Sawyer-Lee R, Gimenez F, Hoogi A, Rubin D. Curated Breast Imaging Subset of Digital Database for Screening Mammography (CBIS-DDSM) [Dataset]. The Cancer Imaging Archive; 2016. 10.7937/K9/TCIA.2016.7O02S9CY.

[12] Suckling J, Parker J, Dance D, et al. Mammographic Image Analysis Society (MIAS) database v1.21 [Dataset]. Apollo – University of Cambridge Repository; 2015. 10.17863/CAM.105113.

[13] Moreira IC, Amaral I, Domingues I, et al. INbreast: toward a full-field digital mammographic database. Acad Radiol. 2012;19:236–248. 10.1016/j.acra.2011.09.014.

[14] Kashiwada Y, Takaya E, Hiroya M, et al. TOMPEI-CMMD Dataset (Version 1) [Dataset]. The Cancer Imaging Archive; 2025. 10.7937/WEZW-BH22.

[15] Strand F. CSAW-CC (mammography) – a dataset for AI research to improve screening, diagnostics and prognostics of breast cancer (Version 1). Karolinska Institutet; 2022. 10.5878/45vmt798.

[16] Nguyen HT, Nguyen HQ, Pham HH, et al. VinDr-Mammo: a large-scale benchmark dataset for computer-aided diagnosis in full-field digital mammography. medRxiv. 2022. 10.1101/2022.03.07.22272009.

[17] Saha R, Lehman CD, Wang S, et al. The EMory BrEast imaging Dataset (EMBED): a racially diverse, granular dataset of 3.4 million screening and diagnostic mammographic images. Radiology: Artificial Intelligence. 2023;5:e220047. 10.1148/ryai.220047.

[18] Logan J, Kennedy PJ, Catchpoole D. A review of the machine-learning datasets in mammography, their adherence to the FAIR principles and the outlook for the future. Sci Data. 2023;10:595. doi:10.1038/s41597-023-02430-6.

[19] Benitez Fuentes JD, Morgan E, de Luna Aguilar A, et al. Global stage distribution of breast cancer at diagnosis: a systematic review and meta-analysis. JAMA Oncol. 2024;10:71–78. doi:10.1001/jamaoncol.2023.4837.

[20] Ozkan I, Yildiz F, Uner S, et al. Stage distribution of breast cancer detected in Turkey’s national mammography screening programme, 2016-2020. Turk J Oncol. 2023;38:159–167. doi:10.5505/tjo.2023.3585.

[21] Ullah F, Kumar K, Rahim T, Khan J, Jung Y. A new hybrid image denoising algorithm using adaptive and modified decision-based filters for enhanced image quality. Sci Rep. 2025;15:8971. doi:10.1038/s41598-025-92283-3.

[22] Bilal A, Alkhathlan A, Kateb FA, Tahir A, Shafiq M, Long H. A quantum-optimized approach for breast cancer detection using SqueezeNet–SVM. Sci Rep. 2025;15:3254. doi:10.1038/s41598-025-86671-y.

[23] Isensee F, Jaeger PF, Kohl SAA, Petersen J, Maier-Hein KH. nnU-Net: a self-configuring method for deep learning-based biomedical image segmentation. Nat Methods. 2021;18(2):203–211. doi:10.1038/s41592-020-01008-z.

[24] Zwanenburg A, Vallières M, Abdalah MA, et al. The Image Biomarker Standardization Initiative: Standardized Quantitative Radiomics for High-Throughput Image-based Phenotyping. Radiology. 2020;295:328–338. doi:10.1148/radiol.2020191145.

[25] Santinha J, Pinto dos Santos D, Laqua F, et al. ESR Essentials: radiomics—practice recommendations by the European Society of Medical Imaging Informatics. Eur Radiol. 2025;35:1122–1132. doi:10.1007/s00330-024-11093-9.

[26] Radzi SFM, Abdul Karim MK, Saripan MI, Rahman MA, Osman NH, Dalah EZ, Noor NM. Impact of Image Contrast Enhancement on Stability of Radiomics Feature Quantification on a 2D Mammogram Radiograph.

[27] Stefano A, Bini F, Giovagnoli E, Dimarco M, Lauciello N, Narbonese D, Pasini G, Marinozzi F, Russo G, D’Angelo I. Comparative Evaluation of Machine Learning-Based Radiomics and Deep Learning for Breast Lesion Classification in Mammography.

[28] Elkorany AS, Elsharkawy ZF. Efficient breast cancer mammograms diagnosis using three deep neural networks and term variance. Sci Rep. 2023;13:2663. doi:10.1038/s41598-023-29875-4.

[29] Sabottke CF, Spieler BM. The Effect of Image Resolution on Deep Learning in Radiography. Radiology: Artificial Intelligence. 2020;2(1):e190015. doi:10.1148/ryai.2019190015.

